# The performance of screening tools for predicting mortality across multi-site international sepsis cohorts

**DOI:** 10.1101/2022.08.27.22279297

**Authors:** Paul W. Blair, Rittal Mehta, Chris Oppong, Tin Som, Emily Ko, Ephraim Tsalik, Josh Chenoweth, Michelle Rozo, Nehkonti Adams, Charmagne Beckett, Christopher W. Woods, Deborah A. Striegel, Mark Salvador, Joost Brandsma, Lauren McKean, Rachael E. Mahle, William Hulsey, Subramaniam Krishnan, Michael Prouty, Andrew Letizia, Anne Fox, Dennis Faix, James V. Lawler, Chris Duplessis, Michael G Gregory, Te Vantha, Alex Owusu-Ofori, Daniel Ansong, George Oduro, Kevin L. Schully, Danielle V. Clark

## Abstract

**Background:** Direct comparisons of sepsis screening tools for prognostication have largely been limited to single-centre or high-income countries despite a disproportionately high burden of sepsis in low- and middle-income countries (LMICs). We evaluated the performance of commonly used sepsis screening tools across prospective sepsis cohorts in the United States, Cambodia, and Ghana.

**Methods:** From 2014 to 2021, participants with 2 or more SIRS (Systemic Inflammatory Response Syndrome) criteria and suspected infection were enrolled in emergency departments and medical wards at hospitals in the Cambodia and Ghana and hospitalized participants with suspected infection were enrolled in the United States. Cox proportional hazards regression was performed, and Harrell’s C-statistic calculated to determine 28-day mortality prediction performance of the qSOFA score ≥2, SIRS score ≥3, NEWS ≥5, MEWS ≥5, or UVA score ≥2, Screening tools were compared to baseline risk (age and sex) with the Wald test.

**Results:** The cohorts included 567 participants (42.9% female) including 187 participants from Kumasi, Ghana, 200 participants from Takeo, Cambodia, and 180 participants from Durham, North Carolina in the United States. The pooled mortality was 16.4% at 28-days. The mortality prediction accuracy increased from baseline risk with the MEWS (C-statistic: 0.63, 95% CI: 0.58, 0.68; p=0.002), NEWS (C-statistic: 0.68; 95% confidence interval [CI]: 0.64, 0.73; p<0.001), qSOFA (C-statistic: 0.70, 95% CI: 0.64, 0.75; p<0.001), UVA score (C-statistic: 0.73, 95% CI: 0.69, 0.78; p<0.001), but not with SIRS (0.60; 95% CI: 0.54, 0.65; p=0.13). Within individual cohorts, only the UVA score in Ghana performed better than baseline risk (C-statistic: 0.77; 95% CI: 0.71, 0.83; p<0.001).

**Conclusions:** Among the cohorts, MEWS, NEWS, qSOFA, and UVA scores performed better than baseline risk, largely driven by accuracy improvements in Ghana, while SIRS scores did not improve prognostication accuracy. Prognostication scores should be validated within the target population prior to clinical use.

**Key questions:** What is already known on this topic – While single-centre cohorts and retrospective analyses have been performed, the optimal sepsis screening tool for prognostication in low- and middle-income countries is unknown.

What this study adds – The MEWS, NEWS, qSOFA scores, but not SIRS, were additive over baseline risk for prognostication in prospective hospitalized infection cohorts, but with variable additive performance within each cohort.

How this study might affect research, practice or policy - Prognostication scores should be validated within the target population prior to clinical use.

## INTRODUCTION

Sepsis, a syndrome resulting from a systemic dysregulated host response to an infection, is estimated to cause six million deaths per year but is likely an underestimate due to limited information from low- and middle-income countries (LMICs) where 87% of the world population live ^1^. Despite declining age-standardized incidence and mortality, sepsis remains a major cause of health loss worldwide and has an especially high health-related burden in LMICs^2^.

Clinical sepsis guidelines developed in the Western world may not be applicable in resource-limited settings and moreover can lead to detrimental effects on sepsis care and management when applied in these conditions due to decreased access to resources to manage iatrogenesis from fluid resuscitation ^3 4^. In contrast to the United States, pathogens that lead to directly lead to vascular injury are common causes of acute febrile illness in Cambodia and Ghana such as dengue virus, malaria, or rickettsia and may alter empiric treatment response ^5^. While early recognition and treatment of sepsis is critical, most sepsis scores or early warning systems were derived from cohorts outside of LMICs. Differences in causes of sepsis, available treatments, and available resources for supportive care should affect management strategies but evidence is limited and optimal clinical scores or biomarkers for sepsis identification are unknown in these settings. Multi-site international sepsis studies are essential for evaluating current and future sepsis tools to ensure effectiveness in resource-limited settings and across populations.

The most validated prognostication scores, SOFA (Sequential Organ Failure Assessment) and the APACHE IV, have been developed for prognostication but require an arterial blood gas and multiple laboratory parameters ^6 7^ that are not widely available in low-resource settings. The qSOFA (quick SOFA) is an abbreviated score that does not require laboratory parameters. The qSOFA is one of the most widely adopted sepsis screening tools and has largely replaced the SIRS (Systemic Inflammatory Response Syndrome) criteria as the standard abbreviated sepsis screening tool as part of the Sepsis-3 definition to identify septic patients ^8^. While the qSOFA and other sepsis screening tools (i.e., Modified Early Warning Score [MEWS], National Early Warning Score [NEWS], and Universal Vital Assessment [UVA]) were developed to identify sepsis, these tools can be used rapidly in the clinical setting and have been studied for their ability to prognosticate mortality among those with suspected sepsis ^9^. Studies have evaluated these tools for predicting in-hospital mortality but the performance of these tools and the prevalence of 28-day mortality, a common metric of sepsis outcomes, have yet to be described across both high- and low-resource settings using similar methods ^9-11^.

We evaluated the performance of sepsis screening tools across prospective multi-site international cohorts that are part of the Austere environments Consortium for Enhanced Sepsis Outcomes (ACESO) consortium ^12^. In contrast to APACHE IV and SOFA, these tools can be quickly performed with limited laboratory test results. We hypothesized that qSOFA may perform poorly in LMIC populations compared to the UVA score due to differences in causes of sepsis. We describe the diverse clinical characteristics, the aetiologies of suspected sepsis within these cohorts, and the performance of sepsis screening tools in current clinical use for predicting mortality at one month post enrolment.

## METHODS

From May 2014 to November 2015, 200 participants were enrolled into a prospective observational study of sepsis at Takeo Provincial Hospital in Takeo Province Cambodia ^13^. This study was followed by a prospective study at Duke University Hospital in Durham, North Carolina, which enrolled 180 participants from December 2014 to March 2016. In Kumasi, Ghana, participants were enrolled at Komfo Anokye Teaching Hospital from July 2016 to October 2017. Study protocols were approved by the Naval Medical Research Center (NMRC) Institutional Review Board (IRB) (Cambodia sepsis study # NMRC.2013.0019; Ghana sepsis study # NMRC.2016.0004-GHA; Duke sepsis study Duke#PRO00054849) in compliance with all applicable Federal regulations governing the protection of human subjects as well as host country IRBs. The study protocol in Cambodia was approved by the Cambodian National Ethics Committee for Health Research (NECHR). The protocol in Ghana was approved by the Committee on Human Research, Publication and Ethics (CHRPE) at Kwame Nkrumah University of Science & Technology. All procedures were in accordance with the ethical standards of the Helsinki Declaration of the World Medical Association. All patients, or their legally authorized representatives, provided written informed consent.

Hospitalized patients ≥ 18 years of age whose attending physician judged them to have an active infection were considered for inclusion for each of the three cohorts. Additional inclusion and exclusion criteria were required in Cambodia and Ghana but not required in the United States protocol. In Cambodia and Ghana, participants were required to meet least two clinical criteria for systemic inflammatory response syndrome (SIRS) during screening. In Cambodia and Ghana, patients were excluded if they had known malignancy, chronic renal/hepatic insufficiency, immunosuppressive conditions (except HIV) or systemic steroid usage that exceeded 20mg/day to prevent confounding in future biomarker studies. Patients were also excluded in Cambodia and Ghana if they had a history of organ transplant, hemodynamically significant gastrointestinal bleeding, anatomic or functional asplenia, acute cardiovascular disease, general anaesthesia, or surgery in the past week prior to enrolment, women who were pregnant, patients who had a haemoglobin less than 7 g/dL or weighed less than 35kg. Hospital physicians who deemed their patients too ill to participate could defer enrolment.

Following informed consent, study team members conducted a detailed medical history, including prior medications, and physical exam. Responses were recorded on a standardized case report form and included demographics, medical history, physical exam findings, and admission diagnoses. Study specific procedures conducted in Cambodia were described in detail by Rozo et al ^14^. Similar enrolment and study procedures were followed in Kumasi, Ghana and in Durham, North Carolina, USA. Blood was collected at the time of enrolment, then at 6 hours later, and at 24 hours later. In Ghana and Cambodia, standardized clinical tests included a peripheral venous blood gas with lactate, complete blood count, complete metabolic panel, optional HIV screening with consent (Alere Determine HIV1/2, Abbott, OK, United States), malaria rapid diagnostic tests (SD Bioline Ag. P.f./Pan, Abbott, OK, United States) and aerobic blood cultures (one aerobic bottle, Bactec 9050, BD, NJ, United States) as part of study procedures in Ghana and Cambodia. Microbiologic results were available if collected through routine clinical care across cohorts. Additional molecular testing and next generation sequencing for pathogens were also performed on blood samples in the Cambodia cohort as previously described ^14^. Participants were followed throughout their hospitalization and a record review performed at discharge. A follow-up interview was performed, and blood samples were collected at a 28-day follow-up visit across cohorts. When patients could not return in person, study team members attempted to conduct an interview with patients or a legally authorized representative by telephone. Fatal outcomes among each discharged participant were also determined. Using clinical data from case report forms and microbiology diagnostic information, clinical adjudication was performed by three physician reviewers (internal medicine or infectious diseases) to determine the source of infection by anatomic location and pathogen class. This was graded on a low, moderate, and high level of confidence by two independent reviewers and a third reviewer served as a tiebreaker for discordant conclusions. If the third reviewer did not agree with either adjudicator, then the decision was determined by committee. Microbiologic results presented include those adjudicated to be clinically relevant to participant’s acute illness.

### Patient and Public Involvement

Patients were not involved in recruitment, design, conduct, or dissemination plans of our research. Results of this study were disseminated to hospital and clinical leadership at Takeo Provincial Hospital and Komfo Anokye Teaching Hospital.

Summary statistics were calculated for the cohorts individually and pooled, comparing baseline demographics (e.g., gender, age, ethnicity, selected medical comorbidities), baseline screening tool scores, physiologic parameters, baseline clinical laboratory values using either Chi-square (categorical values), Fishers exact (categorical values), or Mann-Whitney U tests (continuous values). Prevalence of diagnoses were described for each cohort by organ system and pathogen type (i.e., bacterial, viral, parasitic, or fungal aetiologies) and by anatomic site.

After checking the proportional hazards assumption, Cox regression was performed with bivariable models to evaluate increased risk of death in each cohort by baseline demographics, comorbid conditions, physiologic parameters, and clinical laboratory parameters. Physiologic parameters and clinical laboratory parameters were modelled as dichotomous or ordinal parameters at clinically relevant cut offs. Screening tools were dichotomized according to current usage, including qSOFA score ≥2 (range, 0 [best] to 3 [worst] points), SIRS score ≥ 2 (range, 0 [best] to 4 [worst] points), MEWS ≥5 (range, 0 [best] to 13 [worst] points), NEWS ≥5 (range, 0 [best] to 20 [worst] points), and UVA ≥2 (range, 0 [best] to 13 [worst])^10^ and were evaluated in Cox regression models unadjusted and adjusted for age and sex for risk of death ^9^. Glasgow Coma Scale Score (GCS; range, 3 [worst] to 15 [best] points) of less than 15 was used for estimation of the qSOFA score, and a GCS of ≤3 for unresponsiveness for NEWS, and GCS score 3-15 for the “alert, verbal, pain, unresponsive” scale (AVPU; alert: GCS 13-15, voice: GCS 9-12; pain: GCS 4-8; unresponsiveness: GCS ≤3) score approximation for MEWS ^15 16^. Data was administratively right censored past 28 days. The Harrell’s C-statistic was calculated for each screening tool for each cohort, the cohorts combined, and Cambodia and Ghana cohorts pooled ^17^. This statistic is a performance analogous to area under the receiver operating characteristic curve (AUROC) but accounts for differences over time with survival outcomes. C-statistic confidence interval estimates were evaluated rather than a statistical test due to risk of type 1 error with that approach.^18^ The Cox regression Wald test p-values were calculated for each score covariate was used adjusting for baseline risk estimated by age and sex ^19^. P-values <0.002 were considered significant using a Bonferroni correction for multiple comparisons. Cohort sample sizes were determined a priori through Monte Carlo simulation modelling for prognostic biomarker identification. All statistical analyses were performed in SAS (Statistical Analytical Software, version 9.4), R version 4.0.2 ^20^ or Stata (version 15.0; StataCorp LLC, College Station, TX, USA) ^21^.

## RESULTS

### Summary demographics, sepsis severity, and laboratory findings

There were 567 participants across the cohorts including 187 from Kumasi, Ghana, 200 from Takeo, Cambodia, and 180 from Durham, North Carolina, United States (**Figure 1**). The study population was predominantly male (57.1% male), with more male participants enrolled in Cambodia than at other sites (68.0% vs 55.0% in the U.S. and 52.4% in Ghana). The overall median age was 50 years (interquartile range [IQR], 36 to 63), which was similar across cohorts (**Table 1**). Previously diagnosed comorbid conditions were most common at the U.S. site including a history of cardiovascular (65.6%; N=118), respiratory (42.2%; N=76), or gastrointestinal (36.7%; N=66) conditions.

**Figure 1.**
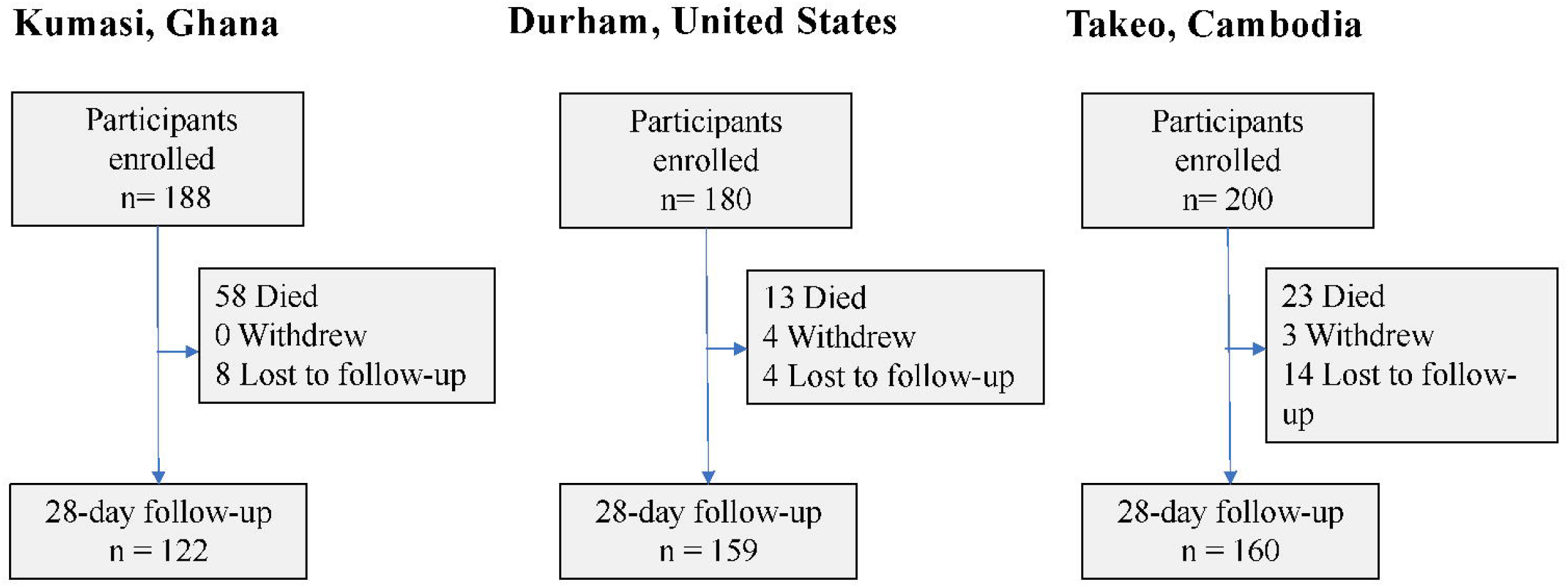
Enrolment flow diagram across cohorts.

**Table 1.** Baseline demographic characteristics stratified by sites.

| **Variable** | **Total** | **Takeo,**<br>**Cambodia**<br>**(n=200)** | **Durham,**<br>**USA**<br>**(n=180)** | **Kumasi,**<br>**Ghana**<br>**(n=187)** |
| --- | --- | --- | --- | --- |
| Female gender – no. (%) | 243 (42.9%) | 64 (32.0%) | 81 (45.0%) | 98 (52.4%) |
| Age – years, median (IQR) | 50 (36 – 63) | 50 (36 – 62) | 52.5 (40 – 63) | 46 (35 – 63) |
| Medical history* – no. (%) |  |  |  |  |
| Cancer | 44 (9.9%) | 0 (0.0%) | 44 (24.4%) | 0 (0.0%) |
| Cardiovascular | 202 (41.4%) | 22 (18.2%) | 118 (65.6%) | 62 (33.2%) |
| Dermatologic | 15 (3.1%) | 1 (0.8%) | 14 (7.8%) | 0 (0.0%) |
| Endocrine | 126 (25.8%) | 6 (5.0%) | 74 (41.1%) | 46 (24.6%) |
| Gastrointestinal | 76 (15.6%) | 4 (3.3%) | 66 (36.7%) | 6 (3.2%) |
| Genitourinary or reproductive | 34 (7.0%) | 1 (0.8%) | 33 (18.3%) | 0 (0.0%) |
| HIV | 26 (4.7%) | 12 (6.2%) | 8 (4.5%) | 6 (3.2%) |
| Neurological | 62 (12.7%) | 1 (0.8%) | 44 (24.4%) | 17 (9.1%) |
| Other | 206 (42.2%) | 48 (39.7%) | 151 (83.9%) | 7 (3.7%) |
| Psychiatric | 143 (29.3%) | 41 (33.9%) | 78 (43.3%) | 24 (12.8%) |
| Renal | 41 (8.4%) | 0 (0.0%) | 41 (22.8%) | 0 (0.0%) |
| Respiratory | 89 (18.2%) | 7 (5.8%) | 76 (42.2%) | 6 (3.2%) |
| Rheumatologic | 29 (5.9%) | 1 (0.8%) | 28 (15.6%) | 0 (0.0%) |
| Surgery | 27 (5.5%) | 0 (0.0%) | 22 (12.2%) | 5 (2.7%) |
| Baseline scores – no. (%) |  |  |  |  |
| MEWS ($\geq 4$) | 315 (57.8%) | 81 (40.7%) | 105 (65.6%) | 129 (69.3%) |
| NEWS score ($\geq 5$) | 324 (61.6%) | 90 (47.9%) | 98 (64.5%) | 136 (73.1%) |
| qSOFA ($\geq 2$) | 139 (25.4%) | 22 (11.1%) | 48 (29.6%) | 69 (37.1%) |
| SIRS ($\geq 2$) | 447 (81.8%) | 125 (68.3%) | 157 (89.2%) | 165 (88.2%) |
| UVA ($\geq 2$) | 199 (37.8%) | 47 (25.8%) | 68 (42.8%) | 84 (45.4%) |
| Baseline scores<br>(median [IQR]) |  |  |  |  |
| MEWS | 4 (3-6) | 3 (2-5) | 1 (0-4) | 1 (1-2) |
| NEWS | 6 (3-8) | 4 (2-7) | 7 (3-9) | 6 (4-8) |
| qSOFA | 1 (1-2) | 1 (0-1) | 1 (0-2) | 1 (1-2) |
| SIRS | 2 (2-3) | 2 (1-3) | 3 (2-3) | 3 (2-3) |
| UVA | 1 (0-3) | 1 (0-2) | 1 (0-4) | 1 (0-4) |

\*There were 79 subjects without comorbidity information in the Cambodia cohort.

Clinical physiologic and laboratory value abnormalities at enrolment were common with median respiratory rate at 24 (IQR: 20 to 30), the median white blood count elevated at 12.05 × 10^9^ cells/L (IQR: 8.13 to 16.6 × 10^9^ cells/L), and median lactate elevated at 2.27 mmol/L (IQR: 1.66 to 3.09 mmol/L) (**Supplementary Table S1**). At enrolment, the proportion of an elevated qSOFA (≥2) at baseline was highest at the Ghana site with 44.4% (N=83) of participants compared to 26.0% (N=52) in Cambodia and 22.2% (N=40) in the United States. The SIRS, MEWS, NEWS, and UVA screening tools were similarly higher in the Ghana cohort.

### Pathogens detected

The most common positive microbiologic results overall included bacteraemia (N=83), respiratory culture growth (N=19), serum hepatitis B surface antigen (N=15), and malaria rapid diagnostic tests (N=11). A minority (121 of 567, 21.3%) of subjects had confirmed infections with complete adjudicator agreement using all available sources of clinical microbiologic results (with the notable addition of RNA sequencing of samples from Cambodia^14^) including 90 (15.9%) bacterial, 17 viral (3.0%), 20 malarial (3.5%), and 2 (0.3%) fungal infections identified across all cohorts (**Supplementary Figure S1**).

In Cambodia, the most common bacterial infections with complete adjudicator agreement were *B*.*pseudomallei* (N=10, with blood or respiratory culture growth), presumptive *M*.*tuberculosis* (N=5, with acid fast positive smears), polymicrobial (N=5), and *O. tsutsugamushi* (N=4, determined by sequencing). The most common causes of bacteraemia (17 total of 200 participants) were *B*.*pseudomallei* (N=8), *E*.*coli* (N=3), and polymicrobial infections (N=3). Three participants had a positive malaria RDT. Fungal infections were uncommon with 1 participant with non-albicans Candidemia and 1 with cryptococcal meningitis. Two individuals had dengue fever (one PCR positive and one adjudicated IgM positive). In Ghana, the most common causes of bacteraemia (culture growth from 28 of 187 participants) were *E. coli* (N=6), *S. aureus* (N=6), *Salmonella spp*. (N=5), and *S. pneumoniae* (N=3). Nine participants had a positive malaria RDT and 15 had a positive hepatitis B surface antigen.

In the United States, the most common causes of bacteraemia (culture growth from 19 of 180 participants) were *E*.*coli* (N=5), *K. pneumoniae* (N=3), polymicrobial (N=2), *Pseudomonas spp*. (N=2), or *S. aureus* (N=2).Viral infections detected by PCR included rhinovirus (N=5), influenza A (N=4), respiratory syncytial virus (N=4), human immunodeficiency virus (N=3), and human metapneumovirus (N=3). There was one participant with *Aspergillus fumigatus* fungal pneumonia.

### Diagnoses and Treatments

Across cohorts, the most common organ system sites of infection were lower respiratory tract infection (28.7%; N=163), multifocal or generalized source of infection (including malaria) (13.6%; N=77), and gastrointestinal (including hepatic) (12.7%; N=72) (**Figure S1a**). The most common antibiotics administered in United States, Ghana, and Cambodia were beta-lactam antibiotics (**Supplementary Figure S2**), but antibiotic regimens varied widely among sites. The most common antibiotics classes used were other antibacterials (e.g., glycopeptide antibiotics, 58.9%), beta-lactam antibacterials, penicillins (51.7%), and cephalosporin and carbapenem antibacterials (44.4%) in the United States, cephalosporins and carbapenems (64.2%), macrolides, lincosamides and streptogramins (37.4%), and other antibacterials (33.7%) in Ghana, and cephalosporins and carbapenems (73.0%), beta-lactam antibacterials, penicillins (46.5%) and aminoglycoside antibacterials (39.0%) in Cambodia.

### Survival

Among all cohorts, 16.4% (N=93) of participants had died at one month, including 58 (31.0%) in Ghana, 22 (11.0%) in Cambodia, and 13 (7.2%) in the U.S (**Figure 1**). Among those that died within one month, median time to death was 4 days (IQR: 1 to 11) in Ghana, 7 days (IQR: 3 to 16) in Cambodia, 10 (IQR: 5 to 19) in the U.S., and 5 days (IQR: 2 to 13) overall. Parameters to calculate the qSOFA score and 28-day mortality were available for 96.4% participants. All screening tools were associated with an increased risk of death (**Figure 2**) with the largest increase among those with an elevated UVA score (**Supplementary Figure S3**). For individuals with a UVA ≥2 there was a 5.45 times increased risk of death (95% CI: 3.39 to 8.76; C-statistic: 0.70) and those with a qSOFA ≥2 had a 4.11 times increased risk of death (95% CI: 2.71 to 6.22; C-statistic: 0.66). Those with an elevated SIRS had a 1.81 times increased risk of death (95% CI: 0.94 to 3.50; C-statistic:0.53). Elevated NEWS (HR: 4.03; 95% CI: 2.24 to 7.26; C-statistic: 0.66) and MEWS (HR: 2.03; 95% CI: 1.28 to 3.23; C-statistic: 0.53) had similarly increased risks **(Figure 3)**.

**Figure 2.**
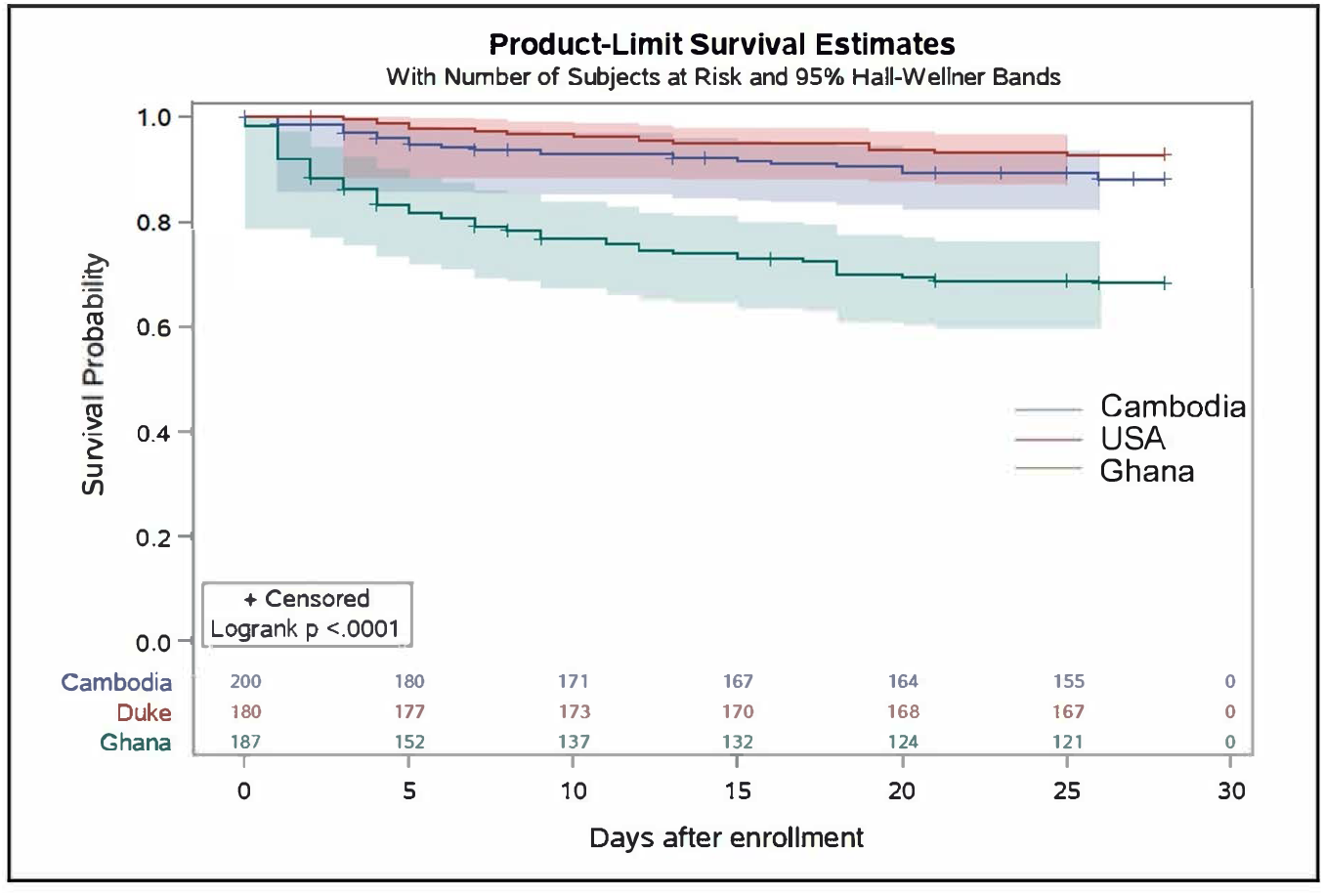
Kaplan-Meier survival plot of 28-day mortality stratified by site.

**Figure 3.**
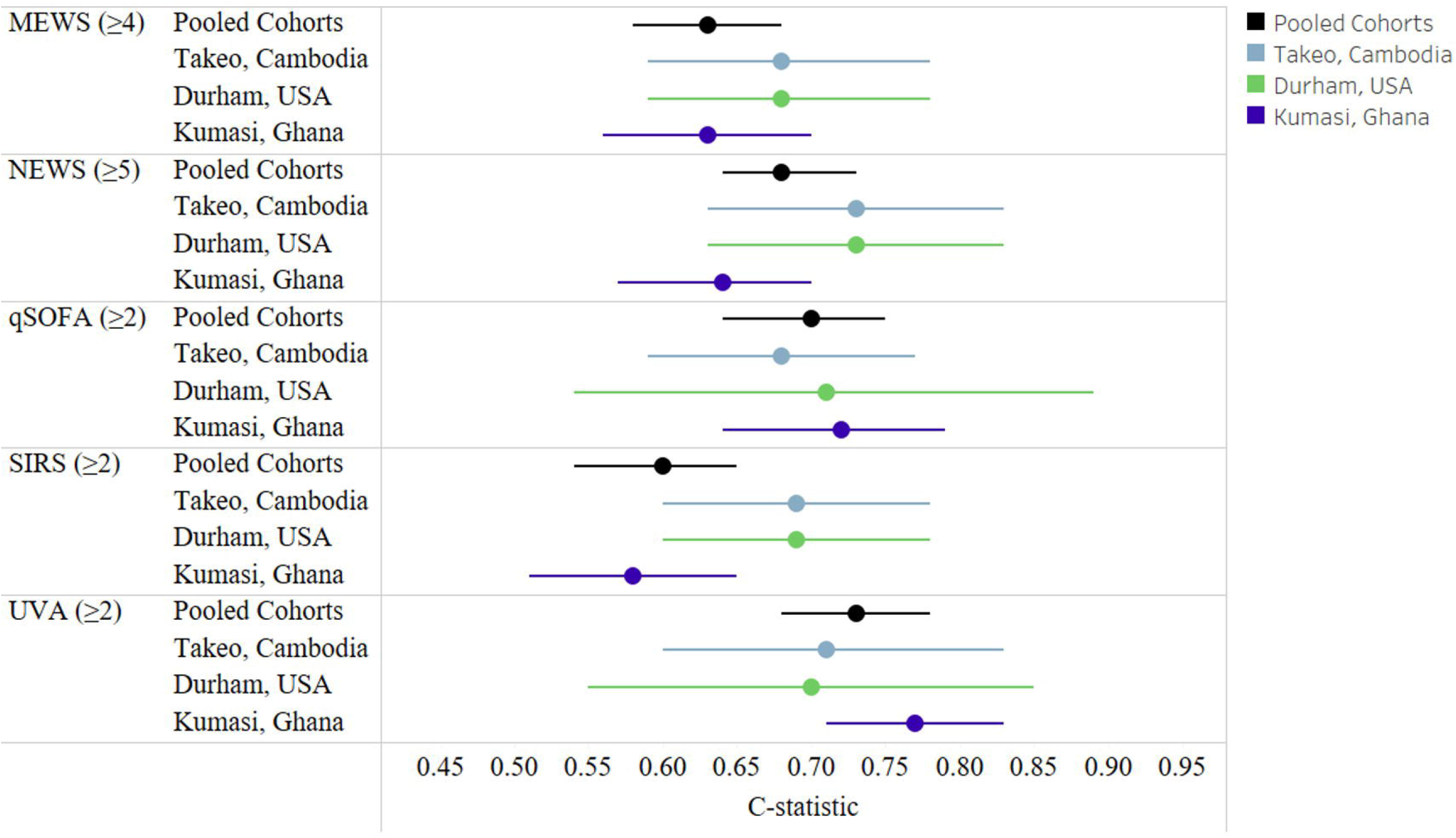
The C-statistic by score overall and by cohort (adjusted for age and sex).

Accuracy in an adjusted Cox model was highest for UVA (0.73; 95% CI 0.68-0.78) followed by qSOFA (C-statistic: 0.70; 95% CI: 0.64 to 0.75) (**Table 2**). The sensitivity for predicting death was highest with SIRS (89%; 95% CI: 80 to 94%) but specificity was lowest (19%; 95% CI: 16 to 26%). The UVA score had a sensitivity of 74% and specificity of 70%. The qSOFA score had the lowest sensitivity (54%; 95% CI: 44 to 65%) but highest specificity (80%; 95% CI: 76 to 84%). We observed that the qSOFA discrimination for mortality was moderate with a C-statistic of 0.70 adjusting for age and sex (**Figure 3**). There was similar qSOFA accuracy in individual cohorts from the United States (C-statistic 0.71; 95% CI: 0.54 to 0.89), Cambodia (C-statistic: 0.68; 95% CI: 0.59 to 0.77), or Ghana (C-statistic: 0.72; 95% CI: 0.64 to 0.79) (**Figure 3**). Similarly, the UVA score had moderate accuracy with a C-statistics on 0.73 (95% CI: 0.68 to 0.78). Other screening scores had similar moderate C-statistic values. The SIRS C-statistic was 0.60 (95% CI: 0.54 to 0.65). Among participants with a NEWS score of ≥5 (62% of the pooled cohort), the C-statistic was 0.68 (95% CI: 0.64 to 0.73) and among those with a MEWS score of ≥4 (58% of the pooled cohort), the C-statistic was 0.63 (95% CI: 0.58 to 0.68) for death. The qSOFA and UVA scores were significantly greater than baseline risk in Ghana in contrast to other scores or cohorts (Table 3). The qSOFA score increased prognostication accuracy in the United States cohort with a p=0.02 but this was not significant after correcting for multiple comparisons. In Cambodia, while not significant after correction, NEWS (p=0.01) and UVA (p=0.01) scores increased accuracy greater than baseline risk.

**Table 2.** Performance characteristics of sepsis score across cohorts for predicting 28-day mortality.

| Score | Sensitivity<br>(95% CI) | Specificity<br>(95% CI) | PPV<br>(95% CI) | NPV<br>(95% CI) | Unadjusted<br>Bivariate<br>Cox model<br>C-statistic<br>(95% CI) | Adjusted*<br>Cox model<br>C-statistic<br>(95% CI) | p-value<br>(Wald test) |
| --- | --- | --- | --- | --- | --- | --- | --- |
| Age and sex | – | – | – | – | – | 0.59<br>(0.53,0.64) |  |
| MEWS$\geq 4$* | 0.73 (0.63,<br>0.82) | 0.45 (0.40,<br>0.49) | 0.21 (0.16,<br>0.26) | 0.89 (0.85,<br>0.93) | 0.59 (0.54,<br>0.63) | 0.63<br>(0.58,0.68) | **0.002\*\*** |
| NEWS$\geq 5$* | 0.86<br>(0.77,0.93) | 0.43<br>(0.38,0.48) | 0.25<br>(0.23,0.28) | 0.93 (0.89,<br>0.95) | 0.65 (0.64,<br>0.67) | 0.68<br>(0.64,0.73) | **<0.001\*\*** |
| qSOFA $\geq 2$* | 0.54 (0.44,<br>0.65) | 0.80 (0.76,<br>0.84) | 0.35 (0.27,<br>0.44) | 0.90 (0.87,<br>0.93) | 0.66 (0.61,<br>0.71) | 0.70<br>(0.64,0.75) | **<0.001\*\*** |
| SIRS $\geq 2$* | 0.89 (0.80,<br>0.94) | 0.19 (0.16,<br>0.23) | 0.17 (0.14,<br>0.21) | 0.90 (0.82,<br>0.95) | 0.53 (0.50,<br>0.57) | 0.60<br>(0.54,0.65) | 0.134 |
| UVA$\geq 2$* | 0.74 (0.64,<br>0.83) | 0.70 (0.65,<br>0.74) | 0.33 (0.27,<br>0.40) | 0.93<br>(0.90,0.95) | 0.70 (0.65,<br>0.74) | 0.73<br>(0.68,0.78) | **<0.001\*\*** |

\*Adjusted Cox model C-statistic is adjusted for age and gender. Note: p-value are from Wald test of the adjusted Cox regression model.
\*\*Significant at p<0.002

**Table 3.** Performance characteristics of sepsis score across cohorts for predicting 28-day mortality stratified by site.

| **Model C-statistic**<br>**(95% CI)** | **Takeo,**<br>**Cambodia** | **p-value** | **Durham, USA** | **p-value** | **Kumasi, Ghana** | **p-value** |
| --- | --- | --- | --- | --- | --- | --- |
| **Age and Sex** | 0.68 (0.59, 0.78) | — | 0.68 (0.54,0.81) | — | 0.57 (0.49, 0.64) | — |
| **MEWS** | 0.68 (0.59, 0.78) | 0.2102 | 0.68 (0.57, 0.79) | 0.4991 | 0.63 (0.56, 0.70) | 0.0097 |
| **NEWS** | 0.73 (0.63, 0.83) | 0.0106 | 0.71 (0.59, 0.84) | 0.2557 | 0.64 (0.57, 0.70) | 0.0022 |
| **qSOFA** | 0.68 (0.59, 0.77) | 0.5101 | 0.71 (0.54, 0.89) | 0.0365 | 0.72 (0.64, 0.79) | **<0.001\*** |
| **SIRS** | 0.69 (0.60, 0.78) | 0.5020 | 0.69 (0.55, 0.83) | 0.5831 | 0.58 (0.51, 0.65) | 0.1882 |
| **UVA** | 0.71 (0.60, 0.83) | 0.0109 | 0.70 (0.55, 0.85) | 0.4753 | 0.77 (0.71,0.83) | **<0.001\*** |

*Note: p-value are from Wald test of the adjusted Cox regression model. Each model is adjusted for age and sex. \*Significant at p<0.002*

When pooling LMIC cohorts (i.e., Ghana and Cambodia), after adjustment for age and sex, the qSOFA (C-statistic: 0.71; 95% CI: 0.66 to 0.77) and UVA scores (C-statistic: 0.76; 95% CI: 0.71 to 0.81) had higher accuracy compared with MEWS (C-statistic: 0.66 (95% CI 0.61 to 0.72), NEWS (C-statistic: 0.70 (95% CI: 0.65 to 0.76), and SIRS (C-statistic: 0.61; 95% CI: 0.55 to 0.67). In contrast, in the United States cohort, NEWS, MEWS, SIRS, qSOFA, and UVA scores after age and sex adjustment each had similar accuracy with C-statistics ranging from 0.66 to 0.71 (**Table 3 and Supplementary Table S3**).

## DISCUSSION

In pooled prospective international cohorts in Cambodia, Ghana, and the United States, the UVA score and Sepsis-3 (qSOFA) performed well with a C-statistic around 0.7 for predicting 28-day mortality. However, this improvement was largely identified in the cohort in Ghana and the accuracy was no different than baseline risk in the Cambodia cohort. There was a trend towards improving prognostication accuracy with the NEWS and UVA score in Cambodia and only with the qSOFA score in the United States. These results suggest that widely used sepsis screening tools may have varying performance for prognostication in diverse settings with different treatment regimens and aetiologies of sepsis. Therefore, screening tools should be selected after validation within populations prior to widespread adoption.

Current sepsis screening tools have had variable performance when applied for prognostication. SOFA or APACHE scores have been developed specifically for prognostication but required parameters including arterial blood oxygen saturation are often not available ^9^. Performance of qSOFA and SIRS for mortality have performed poorly (SIRS, area under the receiving operator curve [AUROC], 0.61; qSOFA: AUROC, 0.61) for prognostication in high-resource settings intensive care unit (ICU) settings ^11^ and in diverse LMICs (adjusted SIRS: AUROC, 0.59; adjusted qSOFA: AUROC, 0.70) ^9^ in prior studies for mortality prognostication. While qSOFA is generally more specific than other screening tools, it is less sensitive than SIRS, MEWS, and NEWS, which is consistent with our data^22^. When applied to sepsis identification, Surviving Sepsis 2021 guidelines recommend against solely using qSOFA, ^23^ due to being a more specific rather than sensitive screening test. Additionally, qSOFA has been found to be inferior to MEWS, and NEWS but more accurate and specific than SIRS for predicting in-hospital mortality and ICU transfer in a large retrospective cohort of over 30 thousand patients in the United States (NEWS: AUROC, 0.77; MEWS: AUROC, 0.73; qSOFA: AUROC, 0.69; SIRS: AUROC, 0.65) ^22^. Different screening scores have been evaluated in prospective cohorts in sub-Saharan Africa (sSA) previously in Tanzania (qSOFA: AUROC, 0.57; MEWS: AUROC, 0.49) ^24^ and Rwanda ^25^ (MEWS: AUROC, 0.69; UVA: AUROC, 0.71; qSOFA: AUROC, 0.65) and in Gabon ^26^ (UVA: AUROC, 0.90; qSOFA: AUROC, 0.77; MEWS: AUROC, 0.72; SIRS: AUROC, 0.70). Given the performance variability that has been previously observed and was observed in this study, it is prudent to evaluate prediction scores within the populations they serve prior to widespread promotion.

The UVA score performed better than baseline risk in the Ghana cohort. Our results externally validated the UVA score for use prognostication of hospitalized patients with suspected sepsis in Kumasi, Ghana and potentially in the region when demographics are similar. The superiority of the UVA score in the Ghana cohort could be related to similarities in infectious causes of illness with other countries in sub-Saharan Africa (sSA) populations from which the UVA score was derived^27^. In contrast to the score derivation study^27^, UVA score performed similarly to qSOFA in Ghana. The accuracy of the UVA scores was not greater than baseline risk in the cohort in Cambodia after adjustment for multiple comparisons. While conclusions may be limited by sample size, sepsis scores derived from the regions of the world with more similar infectious aetiologies may perform better. Our results highlight the importance of validating scores in new patient populations prior to widespread use.

This study had multiple limitations. First, exclusion criteria of immunocompromising conditions except HIV may have led to a skewed populations from Ghana and Cambodia. These exclusion criteria were created to decrease the effect of comorbid conditions or medications on immune biomarkers. However, in Cambodia and Ghana, immunosuppressive medications or diagnoses of chronic liver or kidney disease may be less common in the general population due to limited access to specialists or specialized medications. Additionally, while there were differences in the baseline severity between cohorts, study processes including inclusion criteria were largely standardized across sites improving the comparability of the cohorts in diverse settings. Diagnostic testing differed at each site and mortality specifically due to sepsis could not be determined. Enrolment was by convenience sampling within the referral hospital catchment area and may not be representative of the general population within these countries. Approximation of the mental status for the MEWS scoring using GCS may not be generalizable to the use of GCS at other sites. However, similar MEWS and NEWS performance was observed across sites. Other scores such as the UVA ^25^ have also been evaluated with varying success and future work could be performed to evaluate additional scores. Lastly, due to the limited sample size in each of the cohorts, smaller improvements in accuracy may not have been identified in the Cambodia and United States cohorts that had less deaths compared to the Ghana cohort.

Inexpensive and readily available tools are needed for triage in resource-limited areas in the world to help identify patients that need escalation and possible transfer to higher levels of care. Current widely used sepsis screening tools represent a clinical benchmark for the development of future triage tools. Research is ongoing to assess point-of-care diagnostics within our sepsis cohort research network. Assays with portable and low-cost inflammation biomarkers tests, molecular diagnostics, or point-of-care ultrasound (POCUS) have the potential to augment the performance of clinical screening tools towards a more personalized approach to sepsis recognition and triage.

## CONCLUSION

Sepsis screening tools that are widely used during clinical care had sub-optimal performance for risk stratification in three international cohorts with increased performance of the UVA and qSOFA scores in Ghana compared to baseline risk. There remains a need for reliable, low-cost, and scalable prognostication methods that are validated in diverse settings.

## Supporting information

STROBE checklist

## Data Availability

All data produced in the present study are available upon reasonable request to the authors.

## Funding

Defense Threat Reduction Agency (JSTO-CBA) to Naval Medical Research Center (NMRC) (HDTRA1516108), Defense Health Bureau of Medicine & Surgery to NMRC for Combating Antibiotic Resistance Bacteria (FY1819 0130.1832), Naval Medical Logistics Command Cooperative Agreement (N626451920001).

## Conflicts of Interest

ELT has held equity and consulted for Predigen and Biomeme, and he is an employee of Danaher Diagnostics.

## Disclaimer

K.L.S. is an employee of the US government, and C.B, N.A., C.D., M.P., and A.L. are military service members. This work was prepared as a part of official duties. Title 17 U.S.C. 105 provides that ‘Copyright protection under this title is not available for any work of the United States Government.’ Title 17 U.S.C. 101 defines a U.S. Government work as a work prepared by a military service member or employee of the U.S. Government as part of a person’s official duties. The views expressed reflect the results of research conducted by the authors and do not necessarily reflect the official policy or position of the Department of the Navy, Department of Defense, nor the United States Government.

## Notes

### Author Declarations

Study protocols were approved by the Naval Medical Research Center (NMRC) Institutional Review Board (IRB) (Cambodia sepsis study # NMRC.2013.0019; Ghana sepsis study # NMRC.2016.0004-GHA; Duke sepsis study Duke#PRO00054849) in compliance with all applicable Federal regulations governing the protection of human subjects as well as host country IRBs. The study protocol in Cambodia was approved by the Cambodian National Ethics Committee for Health Research (NECHR). The protocol in Ghana was approved by the Committee on Human Research, Publication and Ethics (CHRPE) at Kwame Nkrumah University of Science & Technology. All procedures were in accordance with the ethical standards of the Helsinki Declaration of the World Medical Association. All patients, or their legally authorized representatives, provided written informed consent.

## References

1. Reinhart K, Daniels R, Kissoon N, et al. Recognizing Sepsis as a Global Health Priority - A WHO Resolution. The New England journal of medicine 2017;377(5):414–17. doi: 10.1056/NEJMp1707170 [published Online First: 2017/06/29]

2. Rudd KE, Johnson SC, Agesa KM, et al. Global, regional, and national sepsis incidence and mortality, 1990-2017: analysis for the Global Burden of Disease Study. Lancet (London, England) 2020;395(10219):200–11. doi: 10.1016/s0140-6736(19)32989-7 [published Online First: 2020/01/20]

3. Maitland K, Kiguli S, Opoka RO, et al. Mortality after fluid bolus in African children with severe infection. The New England journal of medicine 2011;364(26):2483–95. doi: 10.1056/NEJMoa1101549 [published Online First: 2011/05/28]

4. Andrews B, Semler MW, Muchemwa L, et al. Effect of an Early Resuscitation Protocol on In-hospital Mortality Among Adults With Sepsis and Hypotension: A Randomized Clinical Trial. Jama 2017;318(13):1233–40. doi: 10.1001/jama.2017.10913 [published Online First: 2017/10/04]

5. WHO. Dengue: Guidelines for Diagnosis, Treatment, Prevention and Control: New Edition. Geneva 2009.

6. Singer M, Deutschman CS, Seymour CW, et al. The Third International Consensus Definitions for Sepsis and Septic Shock (Sepsis-3). JAMA 2016;315(8):801–10. doi: 10.1001/jama.2016.0287

7. Zimmerman JE, Kramer AA, McNair DS, et al. Acute Physiology and Chronic Health Evaluation (APACHE) IV: hospital mortality assessment for today’s critically ill patients. Critical care medicine 2006;34(5):1297–310. doi: 10.1097/01.Ccm.0000215112.84523.F0 [published Online First: 2006/03/17]

8. Singer M, Deutschman CS, Seymour CW, et al. The Third International Consensus Definitions for Sepsis and Septic Shock (Sepsis-3). Jama 2016;315(8):801–10. doi: 10.1001/jama.2016.0287 [published Online First: 2016/02/24]

9. Rudd KE, Seymour CW, Aluisio AR, et al. Association of the Quick Sequential (Sepsis-Related) Organ Failure Assessment (qSOFA) Score With Excess Hospital Mortality in Adults With Suspected Infection in Low- and Middle-Income Countries. Jama 2018;319(21):2202–11. doi: 10.1001/jama.2018.6229 [published Online First: 2018/05/26]

10. Adegbite BR, Edoa JR, Ndzebe Ndoumba WF, et al. A comparison of different scores for diagnosis and mortality prediction of adults with sepsis in Low-and-Middle-Income Countries: a systematic review and meta-analysis. EClinicalMedicine 2021;42:101184. doi: 10.1016/j.eclinm.2021.101184 [published Online First: 2021/11/13]

11. Raith EP, Udy AA, Bailey M, et al. Prognostic Accuracy of the SOFA Score, SIRS Criteria, and qSOFA Score for In-Hospital Mortality Among Adults With Suspected Infection Admitted to the Intensive Care Unit. Jama 2017;317(3):290–300. doi: 10.1001/jama.2016.20328 [published Online First: 2017/01/24]

12. Krishnan S, Beckett C, Espinosa B, et al. Austere environments Consortium for Enhanced Sepsis Outcomes (ACESO). Shock (Augusta, Ga) 2020;53(3):377–78. doi: 10.1097/shk.0000000000001450 [published Online First: 2019/10/01]

13. Schully KL, Berjohn CM, Prouty AM, et al. Melioidosis in lower provincial Cambodia: A case series from a prospective study of sepsis in Takeo Province. PLoS neglected tropical diseases 2017;11(9):e0005923. doi: 10.1371/journal.pntd.0005923

14. Rozo M, Schully KL, Philipson C, et al. An Observational Study of Sepsis in Takeo Province Cambodia: An in-depth examination of pathogens causing severe infections. PLoS Negl Trop Dis 2020;14(8):e0008381. doi: 10.1371/journal.pntd.0008381 [published Online First: 2020/08/18]

15. Kelly CA, Upex A, Bateman DN. Comparison of consciousness level assessment in the poisoned patient using the alert/verbal/painful/unresponsive scale and the Glasgow Coma Scale. Ann Emerg Med 2004;44(2):108–13. doi: 10.1016/j.annemergmed.2004.03.028 [published Online First: 2004/07/28]

16. Nedea D. AVPU Scale: MDApp; [Available from: https://www.mdapp.co/avpu-scale-calculator-163/ accessed 18 January, 2022.

17. Harrell FE, J., Califf RM, Pryor DB, et al. Evaluating the Yield of Medical Tests. JAMA 1982;247(18):2543–46. doi: 10.1001/jama.1982.03320430047030

18. Han X, Zhang Y, Shao Y. On comparing 2 correlated C indices with censored survival data. Stat Med 2017;36(25):4041–49. doi: 10.1002/sim.7414 [published Online First: 2017/08/02]

19. Pepe MS, Kerr KF, Longton G, et al. Testing for improvement in prediction model performance. Stat Med 2013;32(9):1467–82. doi: 10.1002/sim.5727 [published Online First: 2013/01/09]

20. Team TRDC. R: A language environment for statistical computing. R Foundation for Statistical Computing. 2020

21. StataCorp L. Stata statistical software: release 16 College Station. TX StataCorp LP 2019

22. Churpek MM, Snyder A, Han X, et al. Quick Sepsis-related Organ Failure Assessment, Systemic Inflammatory Response Syndrome, and Early Warning Scores for Detecting Clinical Deterioration in Infected Patients outside the Intensive Care Unit. Am J Respir Crit Care Med 2017;195(7):906–11. doi: 10.1164/rccm.201604-0854OC [published Online First: 2016/09/21]

23. Evans L, Rhodes A, Alhazzani W, et al. Surviving sepsis campaign: international guidelines for management of sepsis and septic shock 2021. Intensive care medicine 2021;47(11):1181–247.

24. Carugati M, Zhang HL, Kilonzo KG, et al. Predicting Mortality for Adolescent and Adult Patients with Fever in Resource-Limited Settings. Am J Trop Med Hyg 2018;99(5):1246–54. doi: 10.4269/ajtmh.17-0682 [published Online First: 2018/09/19]

25. Klinger A, Mueller A, Sutherland T, et al. Predicting mortality in adults with suspected infection in a Rwandan hospital: an evaluation of the adapted MEWS, qSOFA and UVA scores. BMJ Open 2021;11(2):e040361. doi: 10.1136/bmjopen-2020-040361 [published Online First: 2021/02/12]

26. Schmedding M, Adegbite BR, Gould S, et al. A Prospective Comparison of Quick Sequential Organ Failure Assessment, Systemic Inflammatory Response Syndrome Criteria, Universal Vital Assessment, and Modified Early Warning Score to Predict Mortality in Patients with Suspected Infection in Gabon. Am J Trop Med Hyg 2019;100(1):202–08. doi: 10.4269/ajtmh.18-0577 [published Online First: 2018/11/28]

27. Moore CC, Hazard R, Saulters KJ, et al. Derivation and validation of a universal vital assessment (UVA) score: a tool for predicting mortality in adult hospitalised patients in sub-Saharan Africa. BMJ global health 2017;2(2):e000344. doi: 10.1136/bmjgh-2017-000344 [published Online First: 2017/10/31]

