## Supplemental Tables and Figures for "The performance of screening tools for predicting mortality across multi-site international sepsis cohorts"

**Table S1.** Baseline physiologic and clinical laboratory parameters by site at enrollment.

| **Parameter** | **Total**  **Median (IQR)** | **Takeo, Cambodia**  **Median (IQR)** | **Durham, USA**  **Median (IQR)** | **Kumasi, Ghana**  **Median (IQR)** |
| --- | --- | --- | --- | --- |
| **Physiologic parameters** |  |  |  |  |
| Respiratory rate (breaths per minute) | 24 (20, 30) | 24 (20, 28) | 24 (20, 31) | 26 (22, 30) |
| Systolic blood pressure (mmHg) | 120 (100, 130) | 110 (100, 130) | 113 (96, 129) | 127.5 (110, 140) |
| Diastolic blood pressure (mmHg) | 70 (60, 80) | 70 (70, 80) | 64 (56, 75) | 80 (60, 90) |
| Oxygen saturation (%) | 97 (94, 98) | 98 (96, 98) | 95 (92, 97.5) | 97 (95, 98) |
| Temperature (°C) | 37.9 (37, 38.7) | 37.5 (37, 38.5) | 38.1 (36.9, 38.89) | 38.2 (37.4, 38.8) |
| Heart rate (beats per minute) | 105 (94, 118) | 96 (86.5, 105.5) | 111 (99.5, 124) | 111 (99, 118) |
| **Clinical laboratory parameters** |  |  |  |  |
| White blood cells (x10^9^ cells/L) | 12.05 (8.13, 16.6) | 11.9 (8.2, 16.6) | 13.35 (9.7, 17.6) | 10.76 (7.68, 15.41) |
| Platelets (x10^9^ cells/L) | 222 (152.5, 321.5) | 262 (169, 366) | 236.5 (160, 291) | 193 (137, 284) |
| Sodium (mEq/L) | 135 (132, 138) | 135 (131, 138) | 137 (134, 139) | 134 (130, 138) |
| Potassium (mEq/L) | 3.7 (3.3, 4.2) | 3.7 (3.2, 4.1) | 3.9 (3.5, 4.3) | 3.6 (3.2, 4) |
| Sodium Bicarbonate (mmol/L) | 24 (21, 26) | 24 (22, 27) | 25 (22, 27) | 22 (19, 25) |
| Glucose (mg/dL) | 6.56 (5.4, 10) | 6.44 (5.39, 8.28) | 6.69 (5.67, 10.06) | 6.65 (5.2, 12) |
| Blood Urea Nitrogen (mg/dL) | 5 (3.57, 7.9) | 4.29 (3.21, 5.71) | 5.71 (3.57, 10) | 5.4 (3.5, 9.4) |
| Creatinine (mg/dL) | 88.42 (66, 130) | 79.58 (53.05, 88.42) | 106.1 (70.74, 150.31) | 91 (70, 135) |
| Alkaline Phosphatase (U/L) | 86.5 (65, 132) | 98.5 (72, 172) | 80 (63, 106) | 85 (63, 125) |
| Alanine Transaminase (U/L) | 32 (22, 58) | 46 (27, 86) | 22 (18, 40) | 29 (22, 48) |
| Aspartate Aminotransferase (U/L) | 42 (27, 76) | 61 (38, 117) | 29 (21, 45) | 35.5 (25, 65) |
| Bilirubin (mg/dL) | 15 (10.26, 21) | 13.68 (10.26, 20.52) | 15.39 (10.26, 20.52) | 15 (11, 23) |
| Albumin (g/dL) | 3.0 (2.5, 3.5) | 2.9 (2.5, 3.4) | 3.0 (2.5, 3.5) | 3.0 (2.3, 3.6) |
| Total protein (g/dL) | 73 (65, 79) | 74 (68, 79.5) | 67 (57, 72) | 75 (69, 83) |
| Lactate (mmol/L) | 2.27 (1.66, 3.09) | 2.33 (1.79, 3.03) | 1.5 (1, 2.4) | 2.54 (1.8, 3.42) |

**All variables are presented as median, interquartile range*

**Table S2.** Performance characteristics of sepsis score across Cambodia and Ghana sites combined for predicting 28-day mortality.

| **Score** | **Sensitivity (95% CI)** | **Specificity (95% CI)** | **PPV (95% CI)** | **NPV (95% CI)** | **Unadjusted Bivariate**  **Cox model**  **C-statistic (95% CI)** | **Adjusted***  **Cox model**  **C-statistic (95% CI)** | **p-value** |
| --- | --- | --- | --- | --- | --- | --- | --- |
| **Baseline** |  |  |  |  |  | 0.60 (0.54 – 0.66) |  |
| **MEWS ≥4** | 0.74 (0.64 – 0.84) | 0.50 (0.44.3 – 0.56) | 0.28 (0.25 – 0.32) | 0.88 (0.84 – 0.92) | 0.62 (0.57 – 0.66) | 0.66 (0.61 – 0.72) | **<0.001** |
| **NEWS ≥5** | 0.85 (0.75-0.92) | 0.46 (0.38-0.52) | 0.33 (0.29-0.36) | 0.91 (0.85 – 0.94) | 0.65 (0.62 – 0.70) | 0.70 (0.65 – 0.76) | **0.001** |
| **qSOFA ≥2** | 0.54 (0.42 – 0.65) | 0.84 (0.80 – 0.88) | 0.47 (0.39 – 0.55) | 0.87 (0.84 – 0.90) | 0.67 (0.62 – 0.73) | 0.71 (0.66-0.77) | **<0.001** |
| **SIRS ≥2** | 0.88 (0.78 = 0.94) | 0.24 (0.19 (0.30) | 0.23 (0.21 – 24) | 0.89 (0.81 – 0.94) | 0.55 (0.51 – 0.59) | 0.61 (0.55 – 0.67) | 0.066 |
| **UVA ≥2** | 0.75 (0.65 – 0.84) | 0.74 (0.70 – 0.80) | 0.45 (0.40 -0.52) | 0.92 (0.88 -0.94) | 0.73 (0.68 – 0.77) | 0.76 (0.71-0.81) | **<0.001** |

**Table S3.** Performance characteristics of sepsis score across the United States site for predicting 28-day mortality.

| **Score** | **Sensitivity (95% CI)** | **Specificity (95% CI)** | **PPV (95% CI)** | **NPV (95% CI)** | **Unadjusted Bivariate**  **Cox model**  **C-statistic (95% CI)** | **Adjusted***  **Cox model**  **C-statistic (95% CI)** | **p-value** |
| --- | --- | --- | --- | --- | --- | --- | --- |
| **Baseline**  **(age +sex)** |  |  |  |  |  | 0.61 (0.52 – 0.66) |  |
| **MEWS≥4** | 0.60 (0.26-0.87) | 0.33 (0.26 – 0.41) | 0.05 (0.03 – 0.09) | 0.92 (0.85 – 0.96) | 0.53 (0.38 – 0.67) | 0.68 (0.57 – 0.79) | 0.743 |
| **NEWS≥5** | 0.90 (0.56 – 0.99) | 0.37 (0.29 – 0.45) | 0.09 (0.07 – 0.11) | 0.98 (0.89-0.99) | 0.63 (0.54 -0.71) | 0.71 (0.59- 0.84) | 0.256 |
| **qSOFA ≥2** | 0.60 (0.26 -87) | 0.72 (0.65 – 0.79) | 0.13 (0.08 - 0.20 | 0.96 (0.93 – 0.98) | 0.66 (0.51 – 0.81) | 0.71 (0.54 – 0.89) | 0.019 |
| **SIRS ≥2** | 0.92 (0.64 -0.99) | 0.11 (0.07 – 0.16) | 0.08 (0.07 – 0.09) | 0.94 (0.72 – 0.99) | 0.51 (0.45 – 0.58) | 0.66 (0.54 – 0.82) | 0.694 |
| **UVA≥2** | 0.60 (0.26 – 0.88) | 0.58 (0.50 – 0.66) | 0.09 (0.05 -0.14) | 0.95 (0.90 -0.98) | 0.59 (0.44 – 0.73) | 0.70 (0.50 – 0.87) | 0.281 |

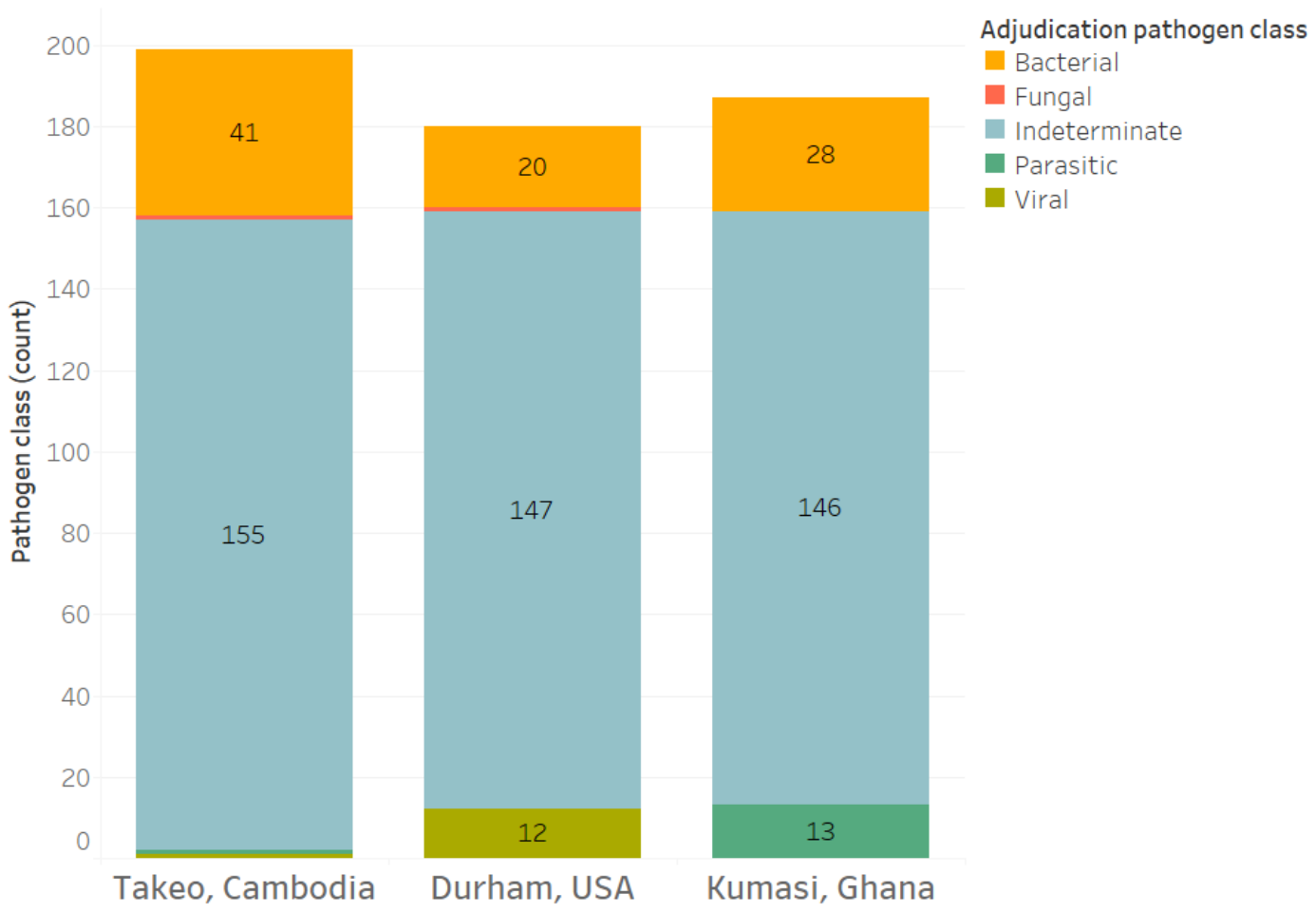

**Supplementary Figure S1.** Distribution of adjudicated pathogen class for each site.

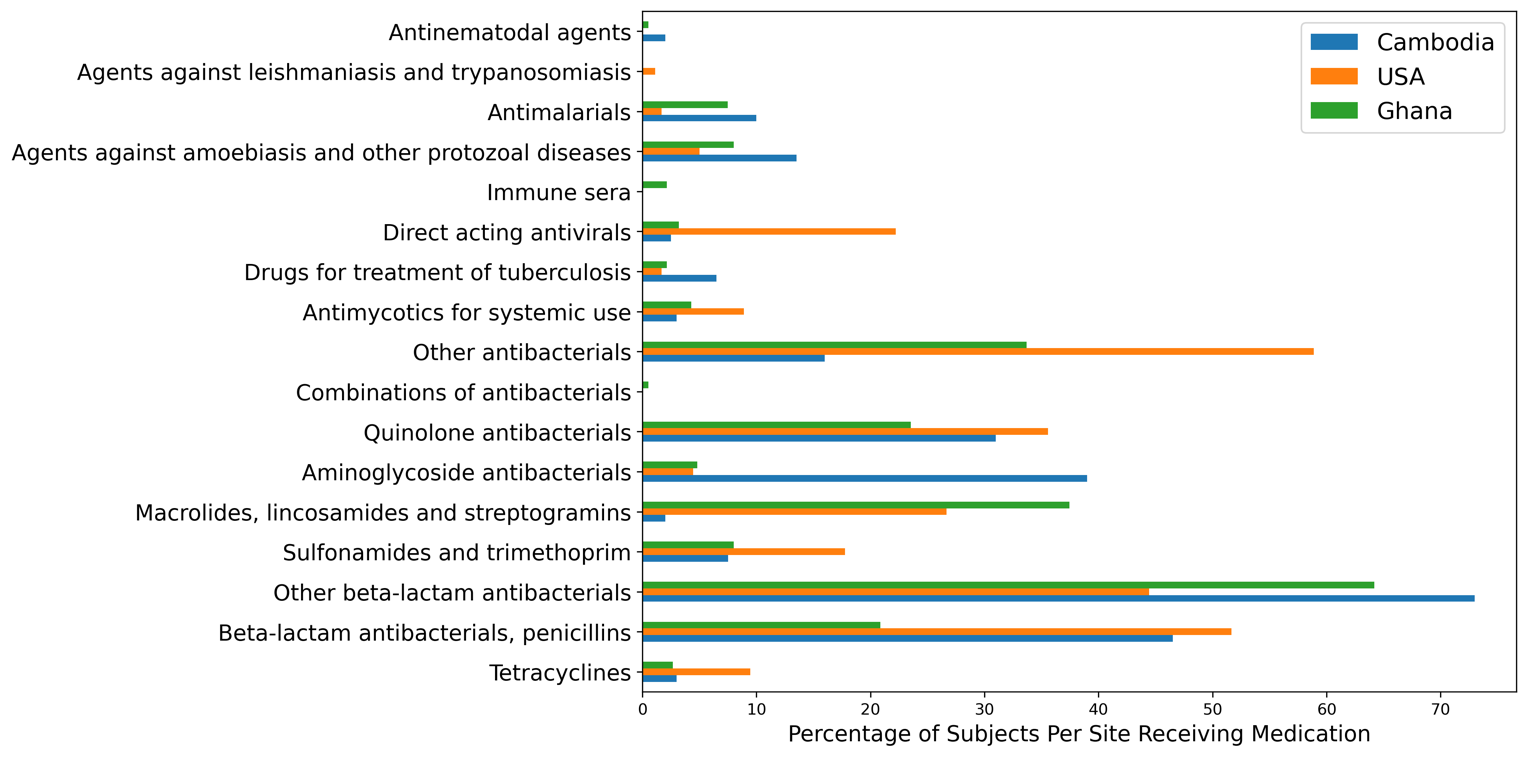

**Supplementary Figure S2.** Prevalence of antibiotics received per site.

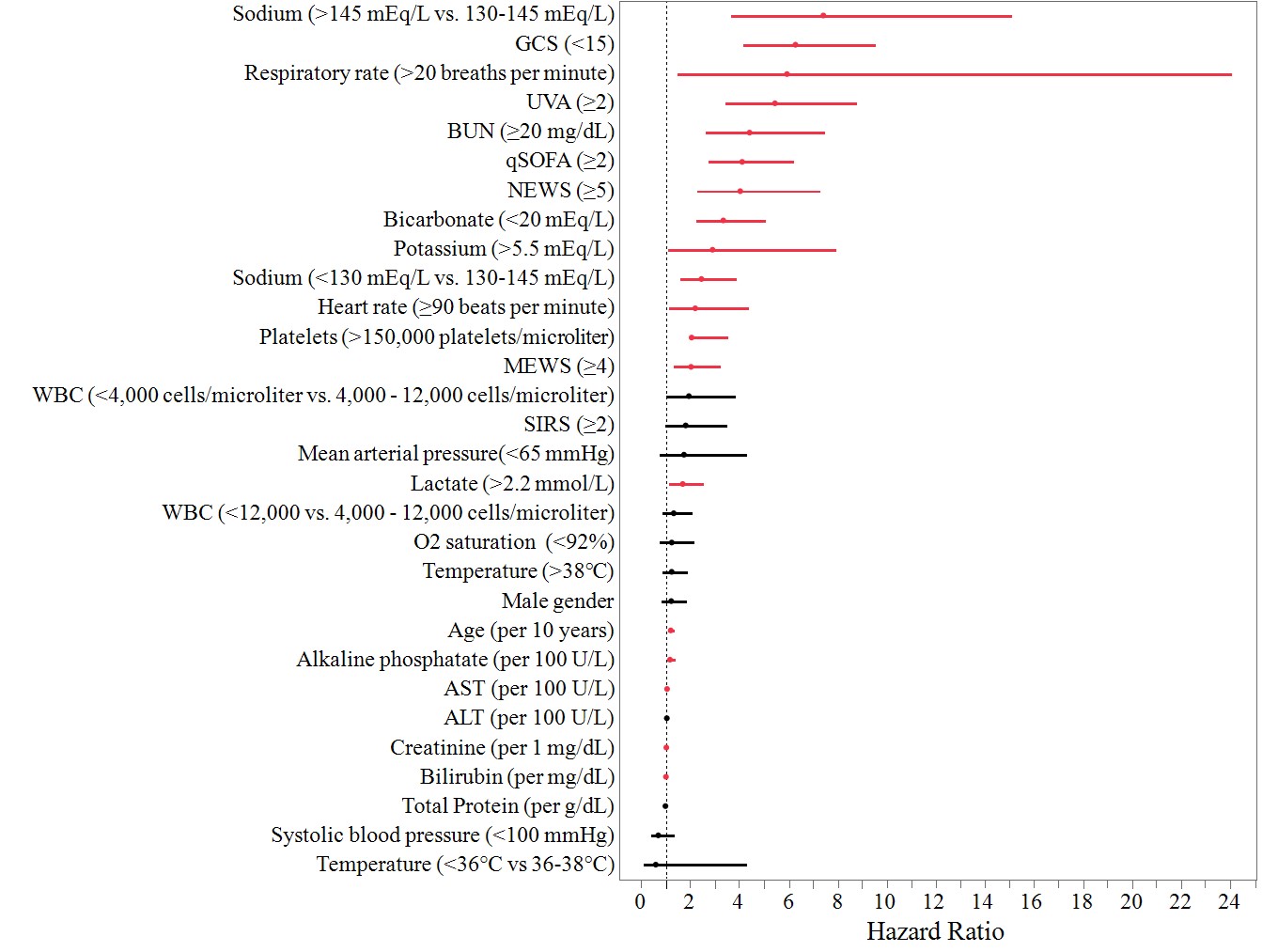

**Supplementary Figure S3.** Forest plot of hazard ratios from bivariate Cox regression models for risk of death at 28-day for sepsis scores, physiologic parameters, and clinical laboratory parameters.
